# Causes of Pediatric Deaths in Lusaka, Zambia: A Quantitative Geographic Information Systems Approach

**DOI:** 10.1101/2024.09.17.24313836

**Authors:** Kristen Sportiello, Mina Shah, Alexandra Buda, Isaiah Mwanza, Manoj Mathews, David R. Bearden

**Author notes:** **Corresponding author:** David R. Bearden.

## Abstract

**Background:** While childhood mortality has been declining in Zambia, it remains high at 58 per 1000 live births. Importantly, many leading causes of mortality in Zambia are preventable. This study was conducted to identify clusters of childhood mortality, causes of death of recently deceased children, barriers to care, and risk factors for mortality in Lusaka, Zambia.

**Methods:** This study was conducted as a prospective cohort study. Family members or lawfully authorized representatives (LARs) were interviewed when they came to pick up death certificates for recently deceased children from Lusaka Children’s Hospital. Each interview included a verbal autopsy, determination of the child’s location of residence, and collection of demographic information. Demographic data was also collected from a healthy control group. Quantitative Geographic Information Systems was used to visualize mortality and evaluate for clustering.

**Results:** Leading primary causes of death included malnutrition (21%), complications of chronic illnesses (16%), and central nervous system infections (13%), while the leading barriers to care were cost (58%) and difficulties with travel (53%). Compared to controls, recently deceased children came from families with significantly lower incomes (1905 Kwacha vs. 2412 Kwacha, p = 0.03) and were significantly more likely to have a history of malnutrition (16.7% vs. 1.4%, p = 0.005). Mortality was clustered in two high-population density, low-income neighborhoods in Lusaka.

**Conclusions:** Systems to reduce financial barriers to care and improve access to transportation could reduce childhood mortality in Lusaka. The aforementioned neighborhoods are ideal locations for public health interventions or improved healthcare services.

## Introduction

While substantial progress has been made over the last 30 years in reducing childhood mortality, rates remain high in most of Sub-Saharan Africa, with under-five mortality rates approximately 14 times higher than those in the United States and Western Europe. Notably, infections and birth complications account for most of this excess mortality. Childhood mortality in Zambia has been decreasing, but it remains high. According to the World Bank, the under-five mortality rate in Zambia was 65 per 1000 live births in 2016 and 58 per 1000 live births in 2019 (1). This high mortality rate is not limited to the most rural parts of Zambia -- the Zambia Statistics Agency’s 2018 Demographic and Health Survey found that the under-five mortality rate in Zambia’s Lusaka Province, which is home to Lusaka, Zambia’s capital city, was 64 per 1000 live births (2). According to the 2020 Vital Statistics Report from the Zambia Statistics Agency and the Ministry of Home Affairs, leading causes of death among Zambian children under 5 include infections specific to the perinatal period and respiratory and cardiovascular disorders specific to the perinatal period, while leading causes of death for Zambian children aged 5-14 include protozoal diseases, human immunodeficiency virus (HIV), and malignant neoplasms (3). Importantly, many leading causes of death among Zambian children are preventable. Zambia has limited resources for public health initiatives, so further research to identify priority areas is needed. In prior research by this group, clustering of both neurocysticercosis and HIV-associated cognitive impairment in high-density, low-income areas in Lusaka was noted, suggesting the feasibility of identifying high-risk neighborhoods for public health interventions (4).

Accurately determining causes of childhood mortality is key to designing effective public health interventions but can be challenging in much of Sub-Saharan Africa. Verbal autopsies (VAs) may be valuable in identifying possible causes of childhood mortality and informing public health policy in settings where medical certification of cause of death is unavailable (5). One study used VAs in Zambia and found that most childhood deaths were from bacterial infection, gastroenteritis, and meningitis and that HIV also played an important role (6). In a univariate analysis, households with lower socioeconomic status were significantly more likely to have had a recent child death (6). This information could be vital to health policy planning. Geographic information system (GIS) technology is also an important tool in evaluation of child mortality. A significant amount of research has been conducted applying GIS technology to diseases and health outcomes worldwide. In Zambia, GIS technology has been applied to the study of malaria, HIV, cholera, and access to healthcare (7–11). Based on these findings, further research using geospatial analysis of various causes of pediatric mortality in Lusaka is of interest.

This study aimed to identify clustering of disease-specific child mortality and child mortality in general in Lusaka, Zambia. Additionally, this study sought to identify nongeographic risk factors for child mortality in Lusaka. We hypothesized that lower income, lack of flush toilets in the home, lack of running water in the home, and history of malnutrition would be associated with child mortality and that mortality would be clustered in high-density, low-income areas. Identification of risk factors for childhood mortality could reveal ideal locations for prevention outreach strategies, identify ideal interventions, and reduce childhood mortality in Lusaka.

## Methods

### Study Design

This was a prospective cross-sectional study with a nested case-control design conducted at Lusaka Children’s Hospital (LCH) at the University Teaching Hospital (UTH) in Zambia. LCH is the largest pediatric hospital in Zambia and a major referral center for the country. All pediatric wards, the emergency department, and the pediatric intensive care unit were surveyed daily Monday through Friday for pediatric deaths by a research nurse or research assistant. The neonatal unit was not included. Data on recently deceased children was collected through comprehensive interviews with family members or lawfully authorized representatives (LARs) who came to pick up children’s death certificates at the hospital. Each interview included a VA administered using the World Health Organization’s VA instrument, which was designed for all age groups and includes questions to assess the context and cause of death (5). Other data collected during each interview included child age, child sex, household income, whether there was a flush toilet in the home, whether there was running water in the home, and location of residence. When internet access was available, interviewees were asked to point out the child’s residence using Google Maps on an iPad, and latitude and longitude data were ascertained using Google Maps. Each interviewee who was unable to circle the child’s location on a map was asked to provide verbal directions to their residence from the hospital, and the study nurse assisted the interviewee in identifying their location of residence.

### Setting/Participants

Interviewees were recruited when they came to pick up death certificates for recently deceased children. The study was described to them, and participants were included in or excluded from the study based on the following criteria. Inclusion criteria included 1) being a family member or LAR of a recently deceased child (younger than 18 years old), 2) coming to UTH to pick up the child’s death certificate, and 3) providing informed consent for study participation. Interviewees were excluded if they were unable to communicate in any language spoken by study staff (English, Nyanja, Bemba, or Tonga), though this did not result in any exclusions. Participants were recruited and interviewed from June 17, 2022 to February 7, 2023. Interviews were conducted at UTH. Medical records of the deceased were reviewed by the study nurse or research assistant and abstracted into a case report form.

Demographic characteristics of recently deceased children were compared to those of healthy control children initially recruited for the HIV-Associated Neurocognitive Disorders in Zambia (HANDZ) study. Full details of the HANDZ study have previously been described (12–14). Briefly, these control children were recruited from the siblings of children attending a general pediatrics clinic. Healthy siblings were recruited instead of children presenting for medical care to reduce the risk of bias. Control participants were aged 8 to 18 years at baseline. Exclusion criteria included history of a central nervous system (CNS) infection such as meningitis or encephalitis and presence of a chronic disease such as diabetes, sickle cell disease, or epilepsy.

### Variables of Interest

Exposures of interest related to mortality were location of the family’s primary residence, family income, presence of a flush toilet in the home, presence of running water in the home, and history of malnutrition. Disease-specific mortality rates were also outcomes of interest, and the primary exposure of interest related to these outcomes was location of the family’s primary residence. The population of each constituency in Lusaka Province, which was used to detect clustering of mortality, came from the Zambia Statistics Agency’s 2022 Census of Population and Housing (15).

### Determination of Cause of Death

Primary and secondary causes of death were determined by consensus between two of the investigators after review of VA and all available records. A single primary cause of death was identified when possible, while any number of secondary contributing factors could be considered. Primary cause of death was determined hierarchically based on the root cause of the admission; for example, in a child who was hospitalized for severe malnutrition before developing a skin infection and sepsis and dying while hospitalized, malnutrition was considered the primary cause of death, while skin infection and sepsis were considered secondary causes. For cases in which consensus could not be reached, or if the consensus was that there was insufficient information to accurately assign cause of death, the cause of death was listed as unknown.

### Statistical Analysis

Statistical analyses were conducted using STATA 17.0 (College Station, TX). Means and standard deviations were calculated for continuous variables, while percentages were calculated for categorical variables. Differences in continuous variables between groups were detected by transforming the variable of interest for normality and then performing a two-sample t-test.

Differences in categorical variables between groups were detected using Fisher’s exact tests. One-sided tests were used as there were pre-established hypotheses regarding the directions of association. Income was controlled for using a multivariable regression model. Listwise deletion was used in regression models, and pairwise deletion was used for all other analyses. For all tests performed, a p-value of 0.05 was considered significant.

### Geographic Analysis

The locations of recently deceased children were used to create a map to visualize mortality in space. First, the latitude and longitude coordinates were imported into a geospatial mapping software program called QGIS (version 3.32) (16). Shapefiles of Zambian constituencies were then added to the map to allow visualization of mortality in space. Clustering of mortality was detected using the observed number of cases of mortality in each constituency in Lusaka Province, the expected number of cases in each constituency in Lusaka Province (determined using the population of each constituency (15)), and a chi-square test.

### Power and Sample Size Calculation

This study was conducted on a convenience sample of all family members and LARs collecting death certificates over the study period. A 2:1 ratio of controls to cases was utilized to improve power. Given the actual number of cases and controls recruited, we had 78% power to detect effect sizes of half a standard deviation or greater when comparing continuous variables between groups.

### Ethics Statement

This study was approved by the institutional review boards of the University of Rochester (protocol #5944), the University of Zambia (reference #1765-2021), and the National Health Research Association of Zambia. Verbal and written informed consent were required from family members or LARs for enrollment in the study.

## Results

### Participants

38 recently deceased children and 75 controls were included in this study. Demographic information for the two groups can be seen in Table 1. Due to differences in inclusion/exclusion criteria between the two groups, controls were significantly older than cases but were otherwise demographically similar except as described below.

**Table 1:** Characteristics of recently deceased children and control participants.

|  | Recently deceased children (n = 38) | Number of recently deceased children missing data for variable | Control participants (n = 75) | Number of control participants missing data for this variable | P-value for test of significant difference (Fisher's exact test or two-sample t-test – see methods for further details) | P-value adjusted for income using multivariable regression model |
| --- | --- | --- | --- | --- | --- | --- |
| Age in years (mean ± standard deviation) | 3.3 ± 4.6 | 0 | 12.7 ± 2.6 | 1 | N/A | N/A |
| Sex (number male, % male) | 19, 50% | 0 | 34, 46% | 1 | 0.70 | N/A |
| Monthly income in Zambian Kwacha (mean ± standard deviation) | 1905 ± 1028 | 12 | 2412 ± 1190 | 25 | **0.03** | N/A |
| Number, % participants with a flush toilet in the home | 14, 37.8% | 1 | 29, 39.7% | 2 | 0.50 | 0.56 |
| Number, % participants with running water in the home | 15, 39.5% | 0 | 39, 54.2% | 3 | 0.10 | 0.39 |
| Number, % participants with a known history of malnutrition (prior to index hospitalization) | 6, 16.7% | 2 | 1, 1.4% | 3 | **0.005** | **0.025** |

### Data on Children’s Deaths

Primary and secondary causes of death are shown in Table 2. A primary cause of death was determined for 27/38 children. It was not possible to determine the primary cause of death for the other 11 children using the information provided. Malnutrition was the leading primary cause of death (8 children -- 21%), complications of chronic illness (e.g. congenital heart disease or cerebral palsy) were the second most common primary cause of death (6 children – 16%), and CNS infection was the third most common primary cause of death (5 children -- 13%). Secondary or contributing causes of death included sepsis (in 31 -- 82%), respiratory infection (in 26 -- 68%), and gastrointestinal infection or CNS infection (in 20 each/53% each). Due to missing data on HIV status in many participants and discordance between parent-reported HIV status and medical records, we were not able to ascertain the role of HIV in childhood mortality in this population. Barriers to care reported by interviewees can be seen in Table 3. The most significant barriers to care were cost (22 children -- 58%) and travel required (20 children -- 53%).

**Table 2:** Causes of death among recently deceased children.

| Cause of death | Number of children for whom this was the primary cause of death | Number of children for whom this was a potential secondary cause of death |
| --- | --- | --- |
| Respiratory infection | 2 (5%) | 26 (68%) |
| Gastrointestinal infection | 1 (3%) | 20 (53%) |
| Central nervous system infection | 5 (13%) | 20 (53%) |
| Sepsis | 0 (0%) | 31 (82%) |
| Malaria | 2 (5%) | 1 (3%) |
| Malnutrition | 8 (21%) | 4 (11%) |
| Stroke | 0 (0%) | 2 (5%) |
| Skin infection | 0 (0%) | 4 (11%) |
| Liver pathology | 0 (0%) | 11 (29%) |
| Malignancy | 0 (0%) | 2 (5%) |
| COVID-19 | 0 (0%) | 3 (8%) |
| Toxic ingestion | 1 (3%) | 0 (0%) |
| Sickle cell | 2 (5%) | 0 (0%) |
| Complications of another chronic illness | 6 (16%) | 4 (11%) |
| Unknown | 11 (29%) | 3 (8%) |

**Table 3:** Barriers to care faced by family/lawfully authorized representatives of deceased children.

| Barrier to care | Number of interviewees |
| --- | --- |
| Cost | 22/38 (58%) |
| Travel required | 20/38 (53%) |
| Family or friends told you not to seek care | 0/38 (0%) |
| Had to remain at home for work/other children | 0/38 (0%) |
| Didn't think it was required | 0/38 (0%) |
| Didn't think doctors could help | 0/38 (0%) |
| Thought it would take too long to get help | 2/38 (5%) |
| Other | 0/38 (0%) |

### Geographic Results

One interviewee was unable to provide sufficient information to determine the child’s location of residence, but it was possible to determine latitude/longitude coordinates for the other 37 children. Locations of recently deceased children can be seen in Figures 1 and 2. As expected, most children who died came from neighborhoods in and around Lusaka, while a small number had been transferred from remote areas (see Figure 1). Among the 24 children who died whose primary residence was within the city of Lusaka (see Figure 2), 63% lived in high-density, low-income areas in the constituencies of Kanyama or Chawama. It was not possible to detect clustering of cause-specific mortality because there was a small number of children representing each cause of death.

**Figure 1:**
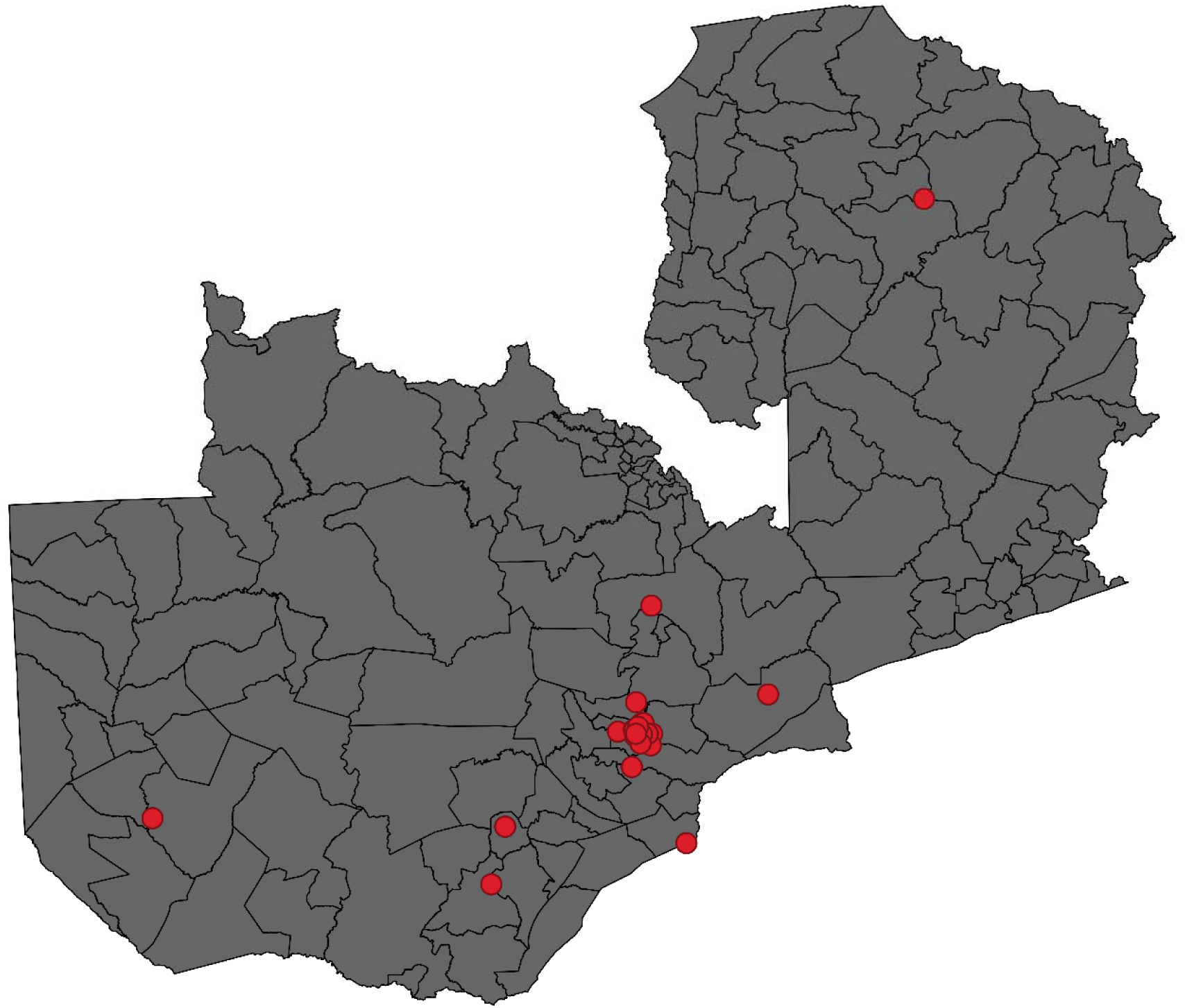
Map of Zambian constituencies with cases of child mortality represented by red dots. The cases are clustered in Lusaka Province, Zambia’s most densely populated province.

**Figure 2:**
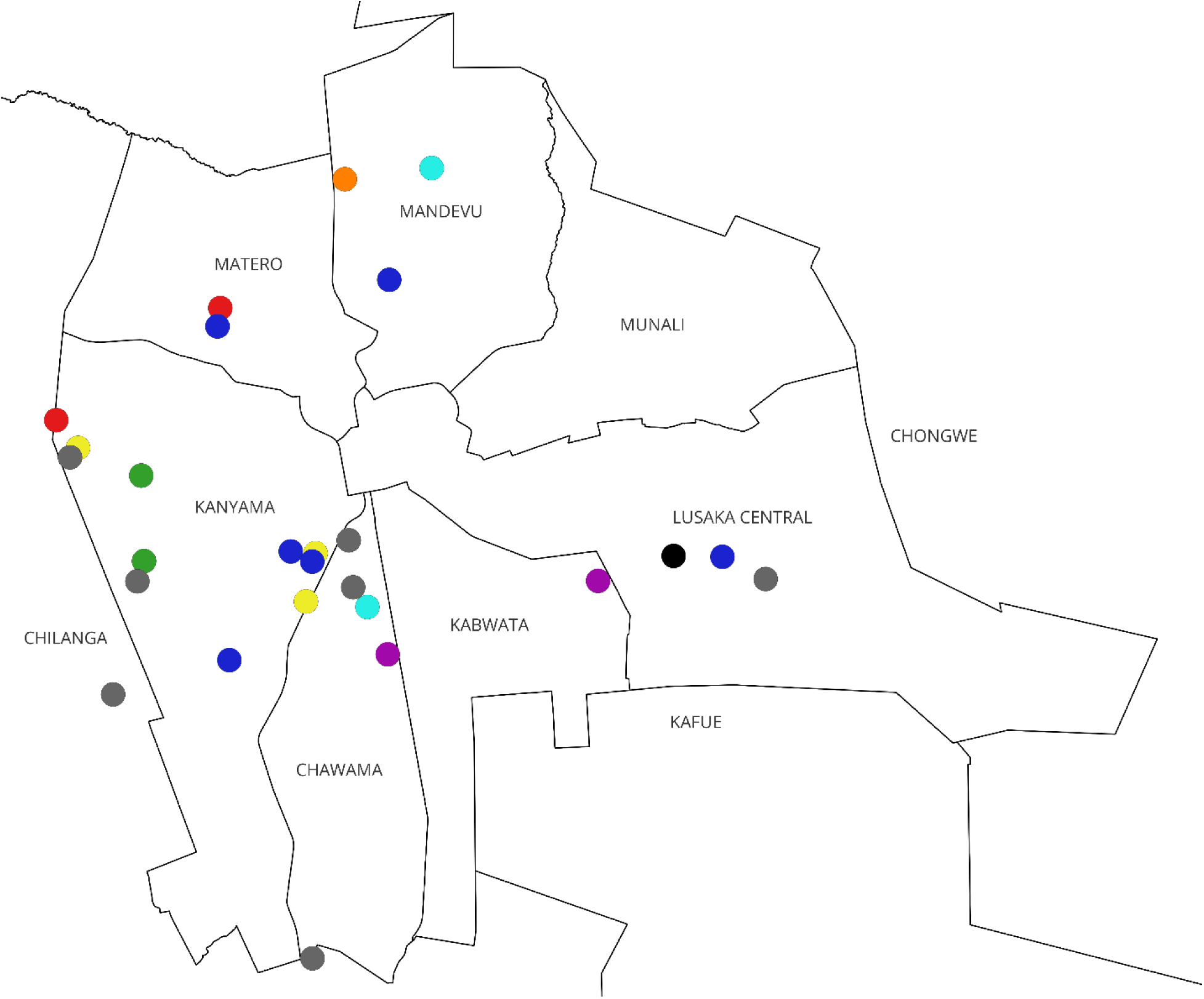
Zoomed-in map of Zambian constituencies illustrating part of Lusaka Province with cases of child mortality represented by dots. Each color represents a different primary cause of death (red: respiratory infection, orange: gastrointestinal infection, yellow: CNS infection, green: malaria, light blue: chronic illness not represented by another color, dark blue: malnutrition, purple: sickle cell disease, black: toxic ingestion, gray: unknown).

### Comparison of Recently Deceased Children to Controls

Data on recently deceased children and controls can be seen in Table 1. There was no significant difference in presence of a flush toilet in the home between the two groups with and without controlling for income. The same was true of running water in the home. Recently deceased children’s families had significantly lower household monthly incomes (1905 ± 1028 vs. 2412 ± 1190 Kwacha, p = 0.03). They were also significantly more likely to have experienced malnutrition (16.7% (6/36) vs. 1.4% (1/72), p = 0.005), even when income was controlled for (p = 0.03).

## Discussion

In this study, we used VAs to identify causes of death among children who died at LCH in Lusaka, Zambia and Quantitative Geographic Information Systems (QGIS) to map cases of childhood death onto neighborhoods in and around Lusaka. This study found that malnutrition, CNS infections, and complications of chronic illness were important drivers of mortality in Lusaka Province. In addition, we identified key barriers to seeking care as perceived cost of care and transportation challenges. The study also found that recently deceased children were significantly more likely to have a history of malnutrition than controls (16.7% vs. 1.4%). Recently deceased children also had significantly lower household monthly incomes (1905 Kwacha vs. 2412 Kwacha). As we had hypothesized, most children who died came from high-density, low-income neighborhoods in Kanyama and Chawama, which are the same neighborhoods in which we detected clustering of neurocysticercosis and HIV-associated cognitive impairment in previous studies. While the study did not detect statistically significant clustering (likely due to small sample size), the maps generated nevertheless suggest target locations for public health interventions. Cluster analysis of disease-specific mortality was not performed as there was a small number of children representing each cause of mortality, but it is notable that the vast majority of deaths in these areas are related to infections or malnutrition and are therefore potentially preventable. Our conclusion that recently deceased children were poorer and more likely to have experienced malnutrition is not surprising, as these are well-established predictors of mortality in low- and middle-income countries (LMICs) (17, 18). It is notable that cost and transportation difficulties remain substantial barriers to care in Zambia, as these represent potentially targetable areas for interventions to improve access to care. Lusaka does not have readily available public sector emergency transportation, and all families in our study used public transportation or private vehicles to bring their children to the hospital, which could have resulted in substantial delays in care.

This study has several similarities to previously reported studies of childhood mortality in the region. Similarly to the World Health Organization (https://files.aho.afro.who.int/afahobckpcontainer/production/files/iAHO_Mortality_Regional-Factsheet.pdf), our study found that noncommunicable diseases now account for a larger percentage of childhood mortality in Zambia. However, our study has some important differences. First, malnutrition and CNS infections accounted for much larger proportions of mortality in our study than what has previously been reported, and lower respiratory infections, malaria, and diarrheal diseases accounted for much smaller proportions. This may be partly due to the methods by which we assigned primary cause of death. That is, if a participant was hospitalized for malnutrition before developing pneumonia and dying, the primary cause of death assigned was malnutrition, while respiratory infection was listed as a secondary cause. However, this may also reflect changing epidemiology in the region, as other studies have also noted significantly less malaria in Lusaka in recent years. Finally, differences in our study may reflect our recruitment strategy, as all participants were hospitalized at UTH. Thus, infants or children who died at home or on their way to the hospital were not represented in our study, which may account for some of the differences between our study and another recent study which used VA to evaluate causes of death in Zambian infants (19).

### Limitations and Risk of Bias

The greatest limitation of this study was the relatively small sample size, which reduced statistical power for cluster analysis of disease-specific mortality. We were able to perform cluster analysis of all-cause mortality, but the small sample size may have prevented the cluster identification. This study was conducted at one referral center in an urban area, so results may not generalize to other areas. Additionally, only children who accessed care at UTH were included in this study, so the results may not generalize to children without access to care. Furthermore, we did not have geographic coordinates for our control group. These coordinates would have allowed us to perform more sophisticated cluster analysis using saTScan software (20, 21). Based on the total reported deaths at LCH over the study period, we enrolled family members/LARs of approximately 10% of all children dying at LCH. Approximately half of families approached for enrollment declined to enroll, and additional families were not enrolled because they did not pick up a death certificate or did so when no study staff was present. Therefore, there may have been meaningful differences between our sample and the total population eligible for enrollment. In addition, we were not able to enroll from the neonatal unit, so this study does not provide insight into mortality related to birth complications. Another major limitation is lack of information on HIV status. While all interviewees were asked about the HIV status of the deceased participant, 100% of interviewees reported the child’s HIV status as negative, which is unlikely in this population. HIV status was often not documented in the medical records, and when present, there was often discordance between the medical records and interviewee report. Future studies in Lusaka will need to ascertain HIV status more accurately.

In conclusion, the combination of VA and QGIS can be a powerful tool for investigating childhood mortality LMICs. We identified two high-density, low-income areas in Lusaka that account for most childhood mortality in Lusaka, and these neighborhoods could be targeted for public health interventions. Systems to reduce financial barriers to care and improve transportation could also reduce child mortality, but further investigation is necessary to identify optimal ways to address these issues. While this study identified some interventions that could be helpful, more studies are required to fully understand child mortality and the best ways to intervene in Lusaka. Detecting clusters of all-cause and cause-specific mortality may be especially helpful, so a study with more participants and geographic coordinates for the control group would be an ideal next step.

## Data Availability

All data produced in the present study are available upon reasonable request to the authors.

## Acknowledgments

This work was supported by the National Institute of Neurological Disorders and Stroke of the National Institutes of Health [K23NS117310] to DB. The authors have no conflicts of interest.

## Data Accessibility Statement

Data is available upon reasonable request by emailing David R. Bearden at

## Notes

### Competing Interest Statement

The authors have declared no competing interest.

### Author Declarations

The institutional review boards of the University of Rochester (protocol #5944), the University of Zambia (reference #1765-2021), and the National Health Research Association of Zambia gave ethical approval for this work.

